# Perspectives on innovation of the diagnostic process in general practice: Q-methodological study

**DOI:** 10.1101/2025.06.04.25328956

**Authors:** Esmée W.P. Vaes, Siamack Sabrkhany, Alma C. van de Pol, Robin E.M. Riphagen, Jochen W.L. Cals, Dorien Zwart, Esther de Groot

## Abstract

**Background:** Amid growing demands, workforce shortages, and rising costs, the evolving role of diagnostics—driven by technological innovation—underscores the urgent need to rethink the diagnostic process with stakeholders at the center. Knowledge on their perspectives on innovation of the diagnostic process is lacking, while it could ensure alignment with their needs and expectations.

**Aim:** To identify perspectives on innovation of the diagnostic process in general practice care.

**Design and setting:** Q-methodological study with stakeholders with different profiles but an informed opinion on the topic.

**Method:** Participants individually sorted 57 statements based on what they thought is most important for innovation of the diagnostic process in general practice care. Statements were collected from literature, media, and group meetings. Factor analyses identified different perspectives, which were subsequently holistically interpreted.

**Results:** We identified five perspectives: 1-innovation through diagnostic transformation, 2-innovation in communication, 3-innovation from a doctor-centered perspective, 4-system reform before innovation: fixing the foundation first, and 5-ambivalence towards innovation. The perspectives differed mostly in their urge for system change and the role for technology in innovation. In some perspectives, changes in the current or future diagnostic system were identified, whereas in others not. Also, the degree to which technology was deemed as the promising way forward differed between the perspectives.

**Conclusion:** This study gives insight into prevailing perspectives of stakeholders on innovation of the diagnostic process in general practice care. Each perspective offers valuable insights, as all are essential for successful innovation of the diagnostic process.

**How this fits in:** Understanding perspectives of stakeholders can help ensure that innovation aligns with the needs and expectations of these stakeholders. This study identified five distinct perspectives of stakeholders on innovation of the diagnostic process in general practice care. We describe why each perspective is important to consider for an innovation to succeed and how shared views can provide common ground for constructive discussions on differences.

## Background

Diagnostics play a crucial role in delivering high-quality healthcare by ensuring patients receive accurate and timely diagnoses and treatment (1, 2). Over the past two decades, innovations in technology and informatics significantly transformed the diagnostic process, elevating its importance in our healthcare system (3). These improvements come with increased usage of diagnostics and interventions, and increased demands on financial resources (4). At the same time, healthcare systems including general practice care must deal with workforce shortages, even as the need for care continues to grow due to an aging population (5). Amidst these challenges, revolutionary technological advancements are being explored for their potential to further innovate the diagnostic process (6). All of this points to a clear need to start thinking about innovation in the diagnostic process, with a focus on stakeholders from the outset.

Many stakeholders are involved in the diagnostic process, working in different settings (i.e. general practice, hospital, diagnostic facility) and fulfilling different roles (applicant, executor, patient, regulator, payer, supplier). It is likely that these stakeholders with different contexts and backgrounds have different interests, concerns and priorities for innovation. Moreover, it is likely that these stakeholders interpret the term “innovation” differently. Innovation is often associated with a new technological tool that we have high hopes for solving our problems. Also, it is still often assumed that once an innovation is proven successful in one setting, it can be implemented and scaled up with minimal obstacles (7). As a result of these high hopes for innovations, they are not critically reviewed for value and relevance and can lead to disappointment when they do not meet their intended results and expectations (7). This shows us that we do not have sufficient understanding of innovation processes.

We argue that it is important to think of innovation not only as the introduction of new tools and technology, but also with organizational, social, and ethical considerations, as previously described by Omachonu and Einspruch (8). Moreover, stakeholder involvement in the innovation process, already from the early onset, is important and inevitable. However, stakeholders were not always sufficiently involved in previous innovation studies (9). It is of special importance to include these different stakeholders when thinking of innovation and to question these stakeholders on their expectations for and visions on diagnostic innovation.

Stakeholders can provide input into setting priorities for innovation of the diagnostic process. It remains unclear what they consider crucial for the future of the diagnostic process. Knowledge of these perspectives could help ensure innovation is aligned with these stakeholders’ needs and expectations (10). Q-methodology is a suitable research method to explore these different perspectives. Its quantitative-qualitative analysis makes it a robust method for studying subjectivity (11).

This study aims to provide insight into the different perspectives of stakeholders in the current debate around innovation of the diagnostic process in general practice care and highlights the importance of understanding these different perspectives.

## Methods

### Study design

Q-methodology combines numerical output (quantitative aspect) for researchers to interpret the meaning of the clustered statements to identify different perspectives (qualitative aspect).

### Data collection

Our data collection consisted of four distinctive steps:

1. Concourse construction

2. Q-sample selection

3. Participant selection

4. Q-sorting and debriefing

#### Step 1: Concourse construction

First, we developed a literature-based theoretical framework to help include all relevant themes in the concourse (**Figure 1**).

**Figure 1.**
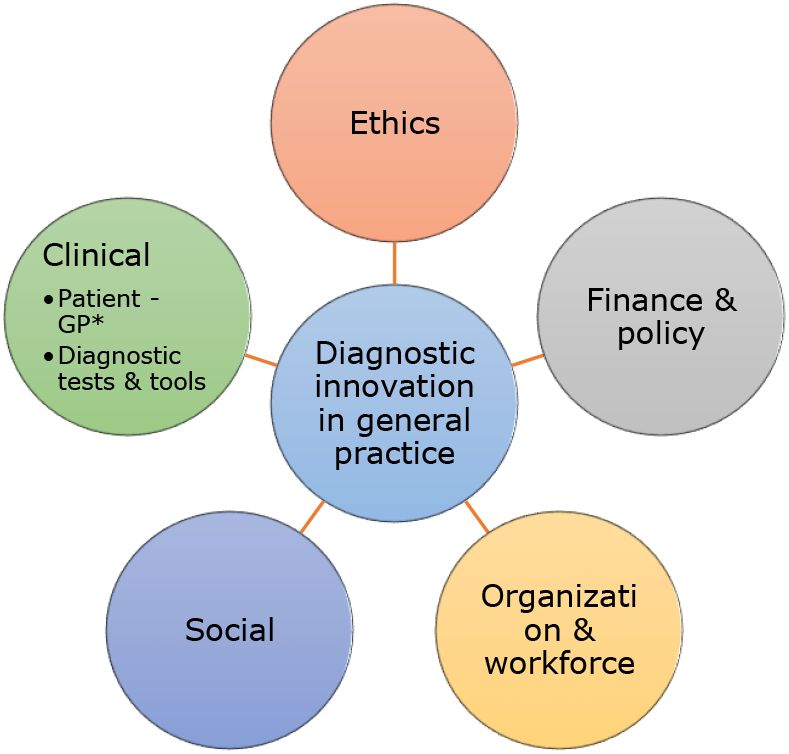
Framework on diagnostic innovation in general practice, including the following themes: ethics, finance & policy, organization & workforce, social, and clinical. During concourse construction, these themes guided the identification of relevant statements concerning diagnostic innovation in general practice. *\*GP = general practitioner*

Second, we performed a literature search in two databases (i.e. PubMed and Embase) and three journals which publish work on healthcare innovation to find recent (2019 to 2023) qualitative studies on the diagnostic process and diagnostic innovation in primary care for quotes. The search terminology and full search strings are presented in **Supplementary Table 1, Supplementary Box 1** and **Supplementary Box 2**. After screening based on title and abstract, articles were included for full-text screening if they described research in primary care setting, were preferably from qualitative nature on diagnosis/diagnostics and/or innovation, and were available in English or Dutch.

Third, we searched Nexis and Google News to add relevant and recent (2019 to 2023) news articles on innovation of the diagnostic process. In addition, a Dutch journal addressing research items for general practitioners (Huisarts & Wetenschap) was searched for relevant and recent (2019 to 2023) articles. The full search strings are presented in **Supplementary Box 3**. Once again, potentially relevant articles were screened to extract insights and opinions.

Fourth, as part of a related study on innovation of the diagnostic process in primary care (12), transcripts of two change laboratory sessions of two separate study groups consisting of general practitioners, patients, medical specialists and policy makers, were read for relevant opinions and quotes on the topic.

In total, we collected 130 statements to form the concourse.

#### Step 2: Q-sample selection

Statements from step 1 were thematically sorted using the framework (**Figure 1**) to ensure comprehensive topic coverage. Duplicates and irrelevant statements were removed by consensus between two researchers (EV and RR). To ensure that the Q-sample included the range and depth of perspectives on the topic, the selection of the final Q-sample statements was done in collaboration with an expert panel, comprising one general practitioner (GP), two GP-researchers, one researcher, and two managers with affinity for primary care diagnostics. These experts combined clinical experiences with theoretical and practical knowledge. Each expert rated the statements on a four-point scale for relevance and clarity for inclusion in the final Q-sample (1 = not essential, 4 = very essential), providing feedback for scores of 2 or lower and suggesting missing statements. For each statement, the content validity index (I-CVI) was calculated (13, 14). Based on the size of the expert panel the I-CVI cut-off point was set at 0.83. Statements below this threshold were revised or removed.

After Q-sample selection by these experts, 59 statements were included in the Q-sample. We organized a pilot session with two researchers (DZ and JC) to evaluate whether statements were understandable and valid enough for meaningful prioritization. Two statements were excluded, resulting in a final Q-sample of 57 statements. A flowchart of the Q-sample selection is shown in **Figure 2**. Statements were presented to participants in Dutch. An English translation using DeepL (15) can be found in **Supplementary Table 2**.

**Figure 2.**
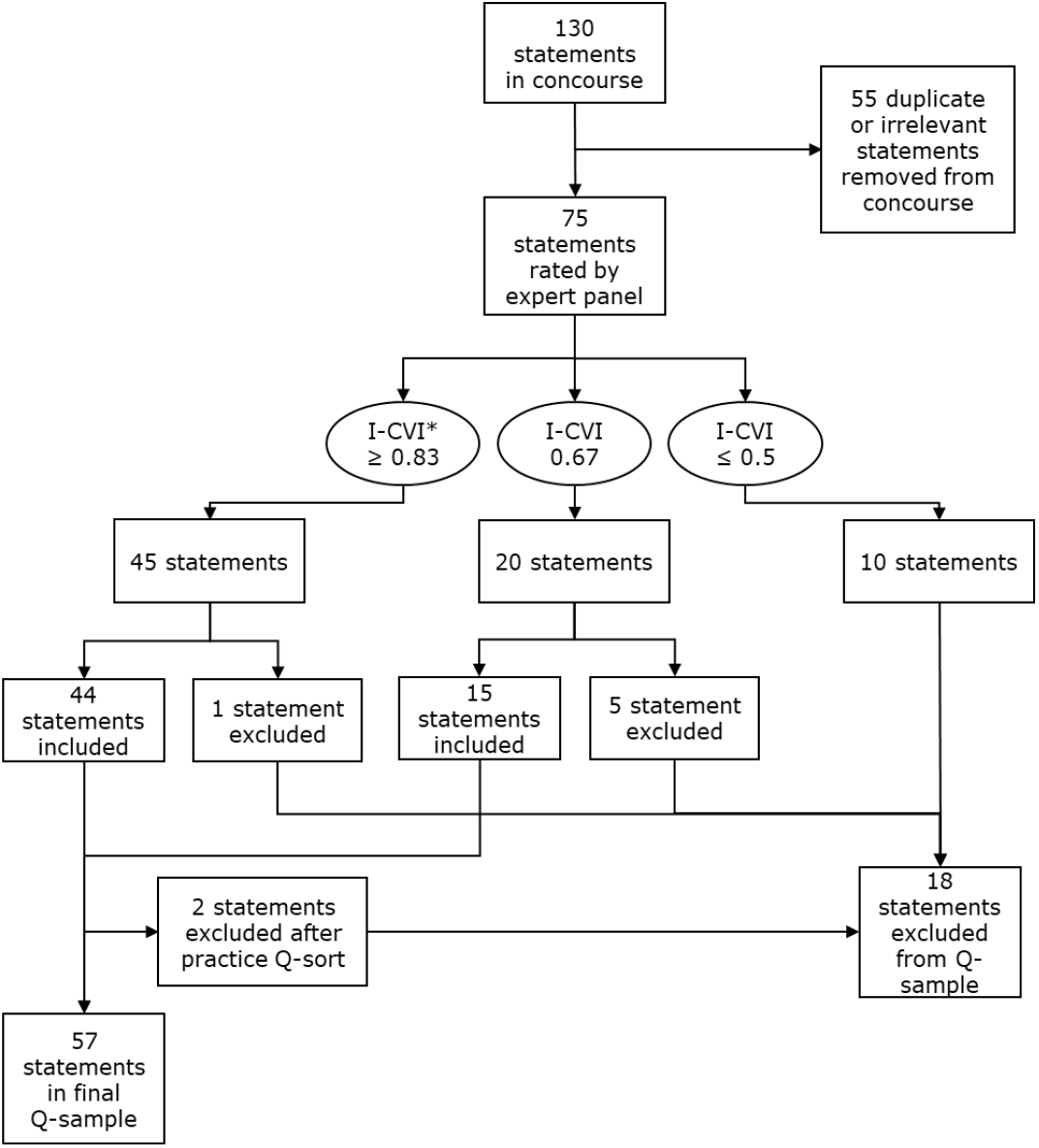
Flowchart of the Q-sample selection. This process included the rating of the statements based on relevance and readability by an expert panel, resulting in 57 statements in the final Q-sample. *\*I-CVI = content validity index*

#### Step 3: Participant selection

We invited participants with an informed opinion on the topic to participate. We considered recruiting participants with one of the following profiles: general practitioners (GPs), GP trainees, patients, nurse practitioners, medical assistants, medical specialists, policy makers, directors/managers, pharmacists, and researchers. We expected that these profiles would have an informed opinion but different views on the topic. We recruited participants on a national scale from within our own network, directly contacting these people and via a social media platform (i.e. LinkedIn). As we focused on innovating diagnostics in general practice, we purposefully invited more GPs and GP trainees than participants with other profiles, as GPs are the primary end-users in this process. In total we included 38 participants in this study.

#### Step 4: Q-sorting and debriefing

Participants were invited for a live or online meeting of approximately one hour with one researcher (EV). After the instructions participants sorted the statements individually.

The researcher was present in case of questions. In case of live meetings participants received the statements and grid on paper. In case of online meetings participants received a link to an online program (https://miro.com/), which allowed participants to drag statements into a box of the grid. The grid is shown in **Figure 3**.

**Figure 3.**
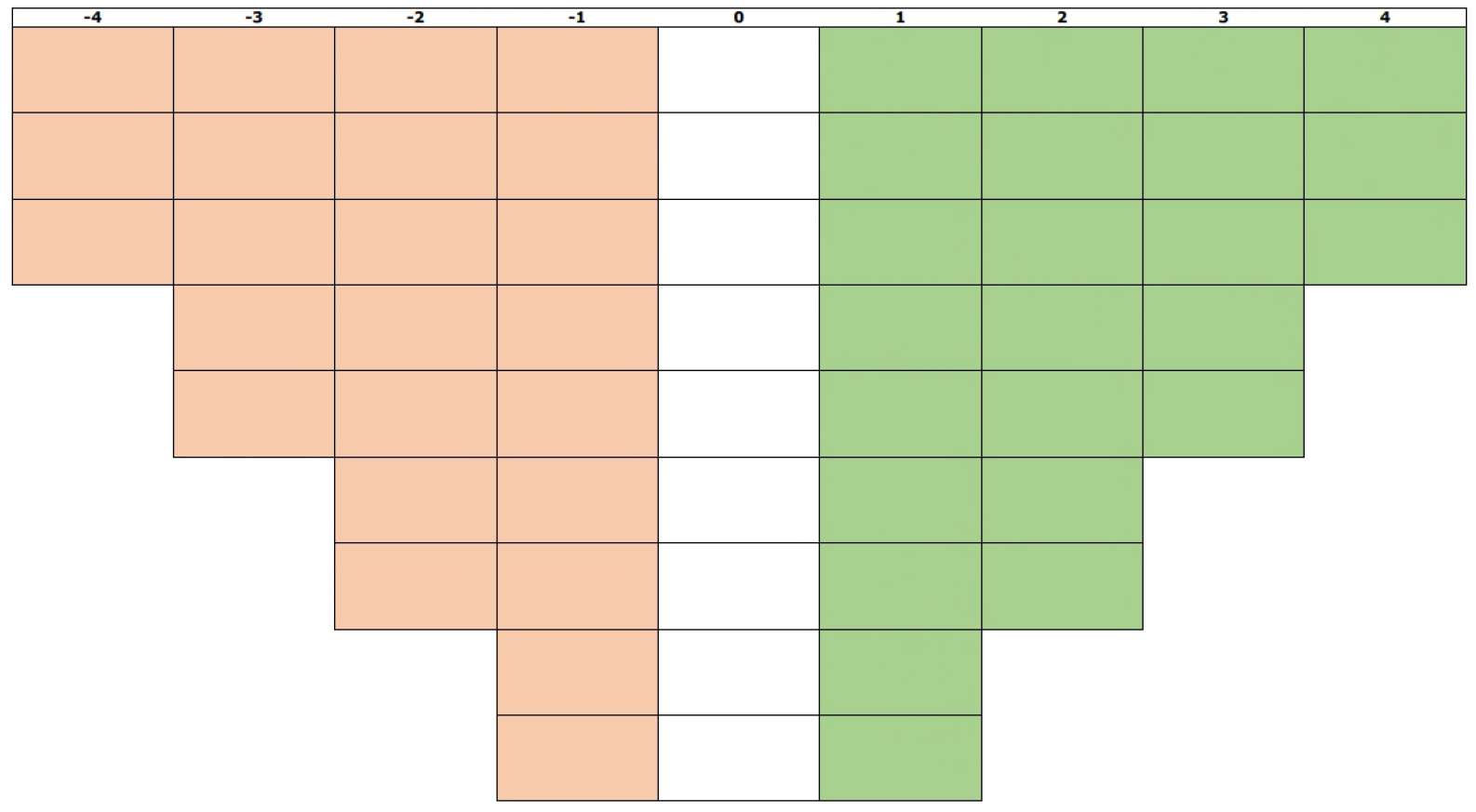
The grid for the sorting of statements. The least important statements were placed in column -4 (in red) and the most important statements in column 4 (in green). All 57 statements were placed in one box in this grid, forcing participants to prioritize these statements.

Participants were given the instruction to place all statements in one box of the grid. To get more familiar with the statements, participants could divide the statements into three stacks: (1) important, (2) neutral, and (3) less important. Then, participants placed the statements onto the grid with a 9-point distribution (−4 to 4). Only a hierarchy existed in the columns, not in the rows of the grid. Participants were forced to make a choice in placing these statements. The Q-sort of one participants was excluded from analysis due to multiple unfilled boxes, indicating either a lack of understanding of the statements or an inability to make a choice. A picture or snapshot was taken of every grid, and the sorting was entered into an Excel sheet.

Immediately after the sorting participants were asked in a short debriefing about the choices they made during the sorting. Participants were also given the opportunity to add additional topics which they missed in the set of statements. All debriefings were audio recorded (live) or video recorded (Teams).

### What do you consider most important in innovation of diagnostics in general practice care?

#### Data analysis

Our data analysis consisted of both quantitative and qualitative steps:

5. Inverted factor analysis

6. Rotation of the factors

7. Factor interpretation

The software tool KADE was used for quantitative analysis (16). One Q-sort was excluded from further analyses because of multiple missing statements in the grid.

#### Step 5: Inverted factor analysis

First an inverted factor analysis was performed to identify which participants sorted the grid in a similar way, shown in a correlation matrix, followed by a centroid factor analysis. A maximum of eight factors were chosen. Multiple ways exist which can be deployed to decide how many factors should be kept (11). Eigenvalues (above 1.0) and study variance were used to determine the number of factors in this study, resulting in a five-factor solution that explained 39% of the total variance.

#### Step 6: Rotation of the factors

The next step was a Varimax rotation, which ensured that every participant that closely resembles a particular perspective was included in that factor. Only Q-sorts with a p-value ≤0.05 were included in a factor.

#### Step 7: Factor interpretation

During an analysis session the research team (EV, SS, AvdP, JC, DZ, EdG) interpreted the holistic sorting of the idealized grid per factor (the sorting of a person who would 100% match within the factor) and used information on statements that distinguished the factors. Per factor the research team wrote down a description of each perspective and placed them in relation to each other on contradicting areas in a conceptual space diagram. If necessary, data from the debriefings were deductively analyzed after this analysis session for extra explanation on each perspective. This also included identifying meaningful quotes to support the description on the perspectives.

### Ethical consent

This study does not fall under the scope of the Dutch Medical Research Involving Human Subjects Act (WMO). It therefore does not require approval from an accredited medical ethics committee in the Netherlands. However, in the UMC Utrecht, an independent quality check has been carried out to ensure compliance with legislation and regulations (regarding Informed Consent procedure, data management, privacy aspects and legal aspects). Participants provided written informed consent before participation in the study.

## Results

Thirty-eight participants performed the sorting of the statements. The Q-sort and data analysis resulted in five different factors representing distinct perspectives on innovation of the diagnostic process in general practice care. After factor development, 29 out of 37 Q-sorts were represented in the selected factors. **Table 1** summarizes the different profiles of the participants that fitted within any of the five factors. All five perspectives included representations of the different profiles.

**Table 1.**
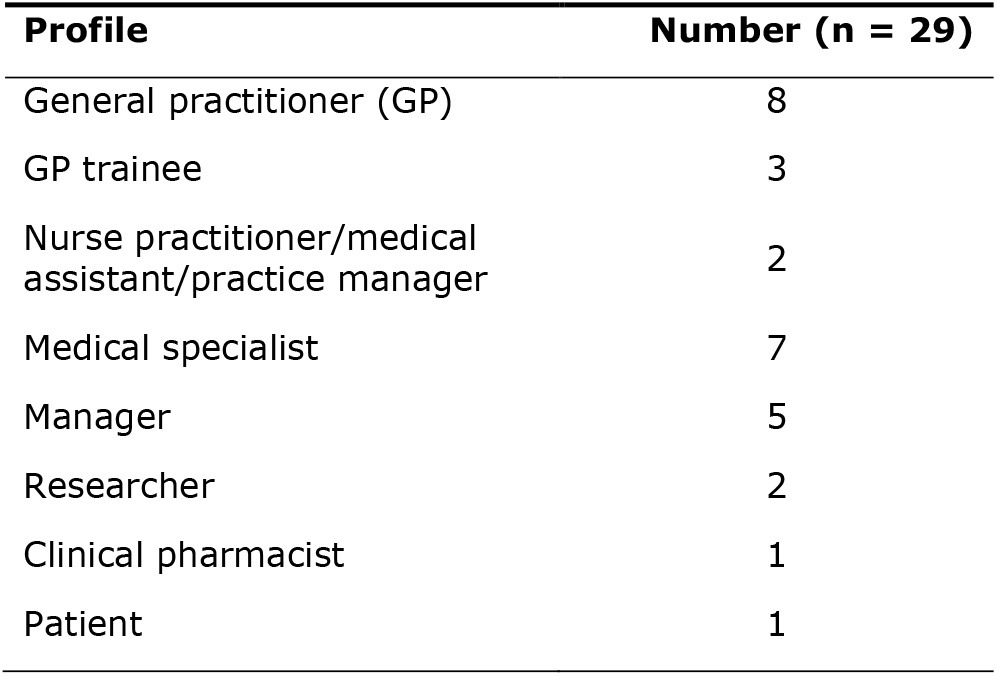
Profile of participants included in the analysis.

### Factor interpretation: five perspectives

The five perspectives on innovation of the diagnostic process were labelled as follows:

(1) innovation through diagnostic transformation;

(2) innovation in communication;

(3) innovation from a doctor-centered perspective;

(4) system reform before innovation: fixing the foundation first;

(5) ambivalence towards innovation.

The perspectives differed most in two areas: ‘dynamics of change’ and ‘the role for technology’. **Figure 4** provides a summary of the five perspectives including a conceptual space diagram. This diagram visualizes the positions of the perspectives in relation to each other regarding these two axes, highlighting not only their differences but also their similarities. **Table 2** shows quotes of participants associated with each perspective. Perspectives varied in the way system changes were thought to be needed for innovation of the diagnostic process, the so-called dynamics of change. Some perspectives thought of system changes as part of innovation of the diagnostic process while others consider system change as a precondition before innovation can take place. Perspectives with a more ‘expansive’ view on system changes thought that transformations in the diagnostic process are needed. Other perspectives had a more preserving, ‘contained’ view on innovation of the diagnostic process. Moreover, the role in which technology was part of the innovation differed between the perspectives.

**Table 2.** Quotes of participants.

| Perspectives | Representative comments |
| --- | --- |
| **1**<br>**Innovation through diagnostic transformation** | • "And the insight of patient records. Yes, in my view that has been for years one of the biggest problems we have and not only the general practitioner, but so also the pharmacy. [...] And with that a lot of duplicate diagnostics are done. With that, a lot of unnecessary diagnostics are done. [...] That creates a huge communication problem, while it is not necessary. And the patient actually assumes that this has been taken care of for a long time." **(P17\*)**<br>• "And the government will also have to play a role, because the more [diagnostics] you do [read: translocate] to the GP, that's a systemic change. Then something will really have to happen that the government also says we support this or not." **(P21)**<br>• "A lot of diagnostic tests can also be done at home, so you send those to the patient, have your patient do them at home and then fill them out digitally." **(P12)** |
| **2**<br>**Innovation in communication** | • "At the end of the day, I think the best interests of the patient should actually be at the forefront of all innovations [...]." **(P14)**<br>• "In my opinion, we also need to get the basics right. And that basis in order is mainly in digital exchange [...] Because then you can [...] do self-testing at home or then you can start monitoring but it starts with getting that data availability in order." **(P6)**<br>• "We are doing a lot of duplicated diagnostics now because certain data is not getting into our systems. [...] That doesn't necessarily have to be a shared record, but to make sure the systems communicate better with each other." **(P13)** |
| **3**<br>**Innovation from a doctor-centered perspective** | • "General practitioners have an enormously broad package they must cover from 'there's not that much going on' to 'immediate action'. I think diagnostics can help with that. But that a doctor can also be supported by technologies to make good choices. That also helps the patient better." **(P37)**<br>• "I think where it [AI] can help me sometimes at least that is with diagnoses where I doubt if I am overlooking a rare cause." **(P28)**<br>• "(Yes, because) all those systems and diagnostics will only work if we as GPs can also think along and provide input for them. And if we do not have a good understanding of how things work, we will never be able to think about them and, as a result, things will be devised that are not at all workable in practice." **(P28)** |
| **4**<br>**System reform before innovation: fixing the foundation first** | • I think that if you put a financial system on it where the cooperation between first and second line [...] the care in the right place or at least the cheapest place from someone who can deliver the care well, [...] then that could be an incentive to cooperate more. **(P27)**<br>• "I think if we start to think that [learning from each other in first and second line] is more normal, that we just solve a problem together instead of continuing to think very strictly in cubicles or in pillars." **(P27)** |
| **5**<br>**Ambivalence towards innovation** | • "I notice that those two things [lagging behind in digitization in healthcare and limited implementation due to high workloads] maintain each other a bit. People say, yes we don't have time at all to invest time in that. And yes, it's just a little bit behind, so to speak." **(P4)**<br>• "I think the workload in a lot of practices is just very high, so yes, there you can look at the patients' interest, but that's not always the interest [...] of the practice." **(P4)** |

\*Participant number

**Figure 4.**
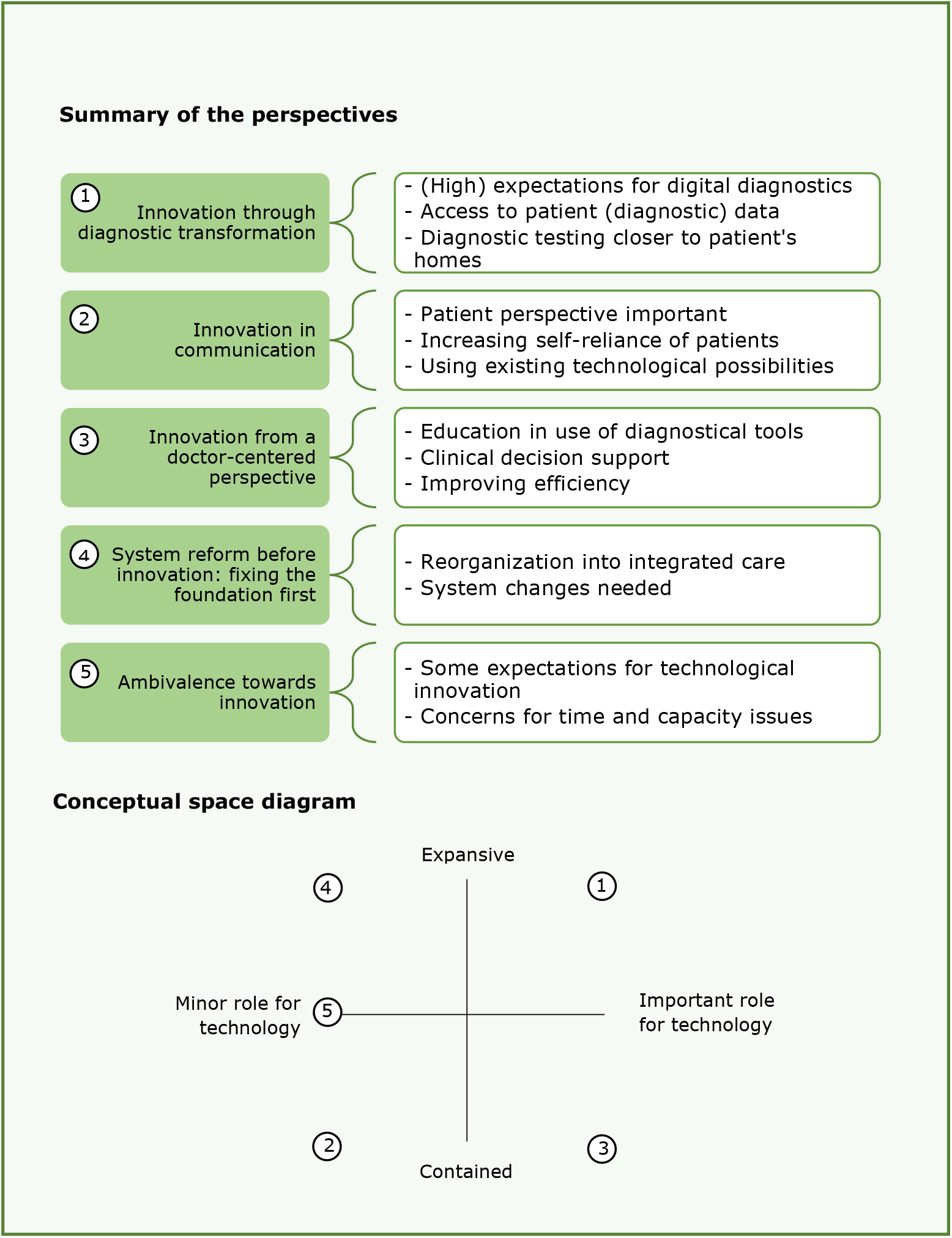
Summary of the five distinct perspectives on innovation of the diagnostic process including a conceptual space diagram depicting the positioning of the different perspectives related to the role for technology in this and the desired level of expansion versus containment. *\*Participant number*

In the descriptions of the perspectives that follows, the numbers between brackets refer to the number of a statement (e.g. #55) and its position on the idealized Q-sort (e.g. - 4), respectively. The idealized grids for each perspective including all 57 Q-sample statements are presented in **Supplementary Table 2**.

### Perspective 1: Innovation through diagnostic transformation

In this perspective, the need for transformation of the diagnostic process is considered urgent. Digital diagnostics is seen as important for ensuring futureproof diagnostics and sustainable healthcare for all patients (#54:4; #55:-4), as well as better patient data visibility and/or accessibility (including diagnostic results) for healthcare professionals in primary and secondary care (#8:3; #7:2). Transformation possibilities in setting and location of diagnostics were also considered important (#27:4; #47:3), e.g., bringing diagnostics closer to the patient, including to the patient’s home (#41:3; #38:-4). To achieve these transformations, a new finance system in healthcare is perceived necessary (#30:4).

In this perspective, system changes as part of the diagnostic process were thought needed with technological and digital innovations playing a vital role in preserving patient care.

### Perspective 2: Innovation in communication

In this perspective, patients fulfill a key role in innovation of the diagnostic process to allow them to become more self-reliant and responsible for their own health and care (#2:3; #33:3). Achieving this requires innovation in communication and data exchange (#7:4; #8:4; #6:3), always regarding the patient’s interests (#56:4). This perspective focused more on improving and implementing existing techniques rather than developing new ones, in which the end-user perspective is highly valued.

In this perspective, the (role of the) patient is central in innovation of the diagnostic process.

### Perspective 3: Innovation from a doctor-centered perspective

In this perspective, innovations that could help GPs in using diagnostics were considered important, like additional education in the use of diagnostic tools (#39:4) and support for GPs in making diagnostic decisions in the consulting room (#3:4), including clinical decision rules (#17:4). Also, improving efficiency of the diagnostic process is also considered important (#48:3), acknowledging a key role for artificial intelligence (AI) herein (#49:3), only if used in a responsible way (#28:3). This perspective did not have concerns about AI limiting patient’s or GP’s autonomy (#14:-4; #57:-4). Finally, this perspective saw opportunity for innovation in the field of funding novel diagnostics (#51:3; #31:2).

In this perspective, innovations of the diagnostic process were mainly centered around doctors, specifically aiming at improving the GP’s work in the consultation room.

### Perspective 4: System reform before innovation: fixing the foundation first

In this perspective, successes of innovation within the current diagnostic system were questioned (#25:3) and suggestions for reorganization into frictionless care were made. To achieve this, several preconditions must be solved before the diagnostic process can be innovated. These preconditions were in collaboration and communication (#6:3), data sharing (#7:4; #8:4), and the finance system of care (#30:4; #53:3). This perspective did not believe in the transformative effects of AI and ICT.

In this perspective, system changes were proposed which are needed first before innovating the diagnostic process.

### Perspective 5: Ambivalence towards innovation

This perspective experienced the current healthcare system as a limiting factor for innovation and was ambivalent towards innovation of the diagnostic process. Although great promises in technological innovation were seen with few ethical concerns (#10:3; #14:-4; #35:-4), lack of time and capacity will prevent innovation to become successful (#42:3).

In this perspective, focus was on constraints that prevent innovation of the diagnostic process rather than suggesting solutions.

## Discussion

### Summary

This study showed that stakeholders have different perspectives on innovation of the diagnostic process in general practice. We identified five perspectives in the current debate around innovation of the diagnostic process in general practice: (1) innovation through diagnostic transformation, (2) innovation in communication, (3) innovation from a doctor-centered perspective, (4) system reform before innovation: fixing the foundation first, and (5) ambivalence towards innovation. The five perspectives particularly differed in their ideas on system change (i.e. contained versus expansive) and the role for technology in innovation (i.e. minor versus important).

### Comparison with existing literature

In perspectives 1 and 4, the importance of transforming the diagnostic process was emphasized while the views differed on the way this transformation should be conducted. In perspective 4, reforming the current healthcare system was emphasized as a prerequisite for any diagnostic innovation. Improvements in interprofessional collaboration, communication, data sharing, and the financial structure of care were seen as necessary first steps before meaningful transformation of the diagnostic process can occur. In contrast, in perspective 1 the belief is reflected that transformation of the diagnostic process is already feasible. For instance, the relocation of diagnostic facilities was identified as a key innovation within this perspective. A Dutch interview study found that general practitioners (GPs) were open for relocating diagnostic care from hospital to general practice, only when also some tasks of the GP could shift to other settings away from the GP. Then, the GP gets time to take over these new tasks from the hospital (17).

Perspectives 2 and 3 were perspectives that considered a lesser need for transformation compared to perspectives 1 and 4. In perspective 2, the focus was primarily on improving communication with patients within the current healthcare system, differing from perspective 4, which prioritized fixing the system first. The emphasis in perspective 2 was less on new technological innovations, in contrast to perspective 3. In this perspective, the emphasis was on technological innovation within the consultation room of the GP, assigning an important role to artificial intelligence, without considering system changes to be necessary. One of the participants quoted (P28, **Table 3**) that she thinks AI-assistance could be valuable in preventing overlooking a rare diagnosis. These opinions correspond to the findings of an interview study by Buck *et al*. (18) in Germany on GPs’ attitudes towards AI-enabled systems. They found that GPs expect that AI-support tools could increase diagnostic quality by helping in diagnosing more accurately and precisely and increase diagnostic efficiency by being able to make rapid diagnosis.

Also in this study, GPs said that they see possibilities for AI in diagnosing rare diseases as an extra support or back-up after making a diagnosis. Concerns were also identified by GPs about the use of AI in the consultation room, for example that it could cause diagnostic bias when GPs rely too much on the suggestions of AI instead of thinking about other diagnosis themselves and neglecting their own medical knowledge and experience. In a study by Razai *et al*. (19) with GPs in the UK, concerns were expressed about the possible increase in unnecessary testing and workload of GPs. Technology intends to rule out any risk, in contrast to the decision making of GPs who always deal with uncertainty. They weigh out the chances of a certain disease against the burden of further diagnostics and treatment, whereas AI will not include any risk and might cause more testing. Therefore, it depends on how AI will be deployed if it indeed benefits diagnostic quality and efficiency (19).

Although our aim was to identify different perspectives, we found that the background of stakeholders did not determine how they thought about innovation of the diagnostic process. Different profiles of stakeholders from different regions in the Netherlands were represented in all five perspectives. This finding indicated that differences in perspective exist between stakeholders with the same profiles (e.g. between GP’s) and from the same region. Similarities in profile and location between stakeholders were no indicators for sharing the same viewpoints.

This study is as far as we are aware of the first to clarify different perspectives on innovation of the diagnostic process in general practice care among stakeholders who are informed and interested in future innovations in this field. Q-methodology enabled us to study subjectivity of different perspectives and priorities. Because this method forced participants to sort statements in relation to each other, it helped to identify contrasting perceptions and opinions (20).

### Strengths and limitations

Q-methodology is commonly used to identify dominant perspectives. Beyond contributing new viewpoints to the literature on innovation of the diagnostic process, this study also describes how these perspectives can be practically applied. We were able to include a diverse group of participants on a national level and identify different prevailing viewpoints without categorizing the participants into different groups. This study applied social science knowledge on innovation processes in thinking about changes in the healthcare system. A strength of the design of our Q-sort study is that we collected and selected our statements comprehensively and transparently. We gathered statements from multiple various sources, including databases, journals, and media, and made use of a self-developed literature-based theoretical framework to make sure that we broadly covered the topic. Moreover, we invited an expert panel for Q-sample selection and recognized the added value to this approach, ensuring a representation of the most important domains in our study. This way, we countered the current critic on Q-methodology that there can be a non-transparent way of concourse construction and Q-sample selection (21).

We recruited participants via purposeful sampling to ensure that participants were stakeholders in the field of the diagnostic process in general practice care. We searched for participants from different backgrounds and contexts, anticipating a range of viewpoints. Based on this type of sampling, it is possible that participants who are more optimistic about innovation of the diagnostic process and thereby willing to participate in this study were more represented in this study. Yet, based on our results, showing different perspectives, we can conclude that we included at least a diverse group of participants. Unfortunately, the analyses of the audio- or video-recorded debriefing data were limited in this study.

### Implications for practice

By reading about the different perspectives stakeholders can recognize themselves in one perspective. Knowledge on the five perspectives can help stakeholders to understand each other’s reactions and attitudes towards innovation, possibly facilitating mutual understanding. Such mutual understanding is particularly valuable for policy makers and funders, as it can support more informed decision-making in policies around the diagnostic process in general practice care. By positioning the different perspectives in the quadrants of the conceptual space diagram presented in **Figure 4**, not only the differences but also the similarities between the perspectives were visualized. Finding these similarities between perspectives could be a good starting point in the discussions about innovation of the diagnostic process. After discussing similarities and establishing this common ground, differences can be further discussed, encouraging stakeholders to look beyond their own perspective.

It is important to realize that these five perspectives exist, and each perspective is valuable to consider, as all perspectives are important for successful innovation of the diagnostic process. If too much focus is on one perspective without considering other perspectives of equal importance, the chances that an innovation succeeds might decrease. To illustrate, although new technological tools are often associated with innovation, this study showed that some perspectives assign a modest role to technology in innovation. These perspectives gave priority to getting the basics right or better alignment with the persons who must undergo diagnostics, the patients, instead of the newest technological innovation. It can help stakeholders who are pro-technology in innovation to get insight that viewpoints on the role for technology can differ. A lack of knowledge and understanding of these perspectives can contribute to failing of some new technological innovations to find their way into practice (10).

## Conclusions

The five identified perspectives of stakeholders on innovation of the diagnostic process in general practice care differed mostly on their ideas on system change and the role for technology in innovation. First discussing similarities between perspectives before highlighting the existing differences could be a good starting point for further conservations on innovation of the diagnostic process. Realizing that each perspective is valuable will increase understanding between stakeholders and increase the success of an innovation.

## Supporting information

Supplementary box 3

Supplementary box 2

Supplementary box 1

## Data Availability

The data that support the findings of this study are available from the authors upon reasonable request and with the permission of UMC Utrecht.

## Declarations

### Ethical approval

This study did not fall under the scope of the Dutch Medical Research Involving Human Subjects Act (WMO), and therefore did not require approval from an accredited medical ethics committee in the Netherlands. An independent quality check has been carried out in the UMC Utrecht (confirmation quality check 23U-0474) to ensure compliance with legislation and regulations. All participants provided written informed consent before participating in this study.

## Acknowledgements

We would like to thank all participants for their contribution to the study and the expert panel who helped with the Q-sample selection.

