## Supplementary table 2 for "Perspectives on innovation of the diagnostic process in general practice: Q-methodological study"

**Table S2** All 57 Q-sample statements and idealized grid for each perspective of stakeholders on innovation of the diagnostic process in general practice care.

| **#** | **Statement** | **Perspective 1** | **Perspective 2** | **Perspective 3** | **Perspective 4** | **Perspective 5** |
| --- | --- | --- | --- | --- | --- | --- |
|  |  | Innovation through diagnostic transformation | Innovation in communication | Innovation from a doctor-centered perspective | System reform before innovation: fixing the foundation first | Ambivalence towards innovation |
| **1** | Digital remote diagnostics is convenient for both patient and caregiver. | 2 | 0 | -1 | 2 | 0 |
| **2** | Additional diagnostics should be used primarily based on the patient's request for help. | -1 | 3 | -2 | -1 | -3 |
| **3** | General practitioners need more support in making decisions about relevant diagnostics. | 0 | -2 | 4 | 1 | -1 |
| **4** | The more diagnostic tests are feasible in general practice itself, the better. | -2 | -4 | -1 | -3 | -4 |
| **5** | Doctors can make better diagnostic decisions using Artificial Intelligence. | 1 | 0 | 1 | 2 | 3 |
| **6** | Better communication between general practitioners and medical specialists is necessary to improve the diagnostic process. | 1 | 3 | 0 | 3 | -1 |
| **7** | Patient records must be immediately accessible to primary and secondary care providers. | 2 | 4 | 2 | 4 | 3 |
| **8** | A single national electronic patient record must be developed. | 3 | 4 | 1 | 4 | 2 |
| **9** | There must be more and larger health centers with diagnostic facilities in which general practitioners and specialists work together. | -1 | -1 | -3 | 3 | -2 |
| **10** | The use of Artificial Intelligence systems is a solution to capacity problems in healthcare. | 3 | 0 | -4 | -2 | 3 |
| **11** | There should be more focus on implementing diagnostic innovations in practice rather than on initial development. | -2 | 2 | 0 | 0 | 1 |
| **12** | Clearer agreements must be made on the use of reliable home measurement devices. | 2 | 0 | 1 | 2 | 2 |
| **13** | Regional agreements should be made between general practitioners, hospitals and laboratories on diagnosis requests to avoid double diagnosis. | 1 | 1 | -1 | 0 | 2 |
| **14** | The use of Artificial Intelligence in diagnostic decisions limits the patient's freedom of choice. | -3 | -3 | -4 | -3 | -4 |
| **15** | The patient should have a greater role in deciding whether or not to use diagnostics. | -3 | 1 | -3 | -1 | -2 |
| **16*** | Primary care physicians are reluctant to embrace innovation in diagnostics. | -2 | -2 | -1 | -2 | 0 |
| **17** | Clinical decision rules are useful in determining whether an indication exists for requesting additional diagnostics. | 3 | 3 | 4 | 3 | -1 |
| **18** | The use of ultrasound in general practice improves the diagnostic process. | -1 | -1 | 0 | -2 | -2 |
| **19** | Diagnostic tests in general practice are preferably rapid tests. | 2 | 2 | 0 | -4 | 3 |
| **20** | Decision scores and models perform better in making diagnostic decisions than GPs. | 1 | -4 | -1 | -1 | -2 |
| **21** | Healthcare is far behind in digitization compared to other sectors. | -2 | 0 | 0 | -1 | 4 |
| **22** | Digitization of care and diagnostics is moving faster than the current healthcare system can handle. | 0 | -2 | -1 | -1 | 1 |
| **23** | Healthcare providers should receive education aimed at dealing with Artificial Intelligence systems. | 0 | 1 | 2 | 0 | 4 |
| **24** | More attention should be paid to the professionalization of general practitioners with regard to dealing with technological developments. | -1 | -1 | 1 | -1 | 1 |
| **25** | The fragmentation of healthcare hinders the development of innovations in diagnostics. | -1 | 2 | -2 | 3 | 2 |
| **26** | The results of rapid tests performed in the general practice should be automatically entered into the hospital record. | 2 | 3 | 1 | 1 | 0 |
| **27** | Innovation of the diagnostic process should aim to move low complex second-line care to primary care. | 4 | -4 | 1 | -1 | -3 |
| **28** | The responsible use of Artificial Intelligence systems should be seen as a medical skill and should therefore be part of physician training. | 1 | 2 | 3 | 0 | 2 |
| **29** | A quality mark should be established for diagnostic innovations in family medicine. | -2 | -3 | 2 | -2 | 0 |
| **30** | A new funding system that is more focused on collaboration between primary and secondary care should be developed. | 4 | -3 | -2 | 4 | -1 |
| **31** | The government should be less reluctant to invest in diagnostic innovation. | -3 | 0 | 2 | 0 | -2 |
| **32** | Digital remote diagnostics should be more accessible to vulnerable groups. | 2 | -1 | -1 | 2 | -2 |
| **33** | A central digital healthcare platform gives patients more control over their own health. | 0 | 3 | -3 | -3 | -2 |
| **34** | Test results should be fed back to patients in understandable language. | 1 | 2 | 3 | 1 | 2 |
| **35** | The use of Artificial Intelligence in the consulting room is not good for the doctor-patient relationship. | -4 | -2 | -3 | 1 | -4 |
| **36*** | More rapid point-of-care tests should be available for use during a visit. | 0 | 1 | 0 | 0 | 1 |
| **37** | Deploying Artificial Intelligence-based systems speeds up and improves the diagnostic process. | 2 | -1 | 1 | 0 | 0 |
| **38** | Home measurements by patients are not reliable enough as diagnostics. | -4 | -3 | -1 | -4 | -3 |
| **39** | GPs should be better trained in the use of diagnostic tools. | -2 | -1 | 3 | 1 | 2 |
| **40** | The use of Artificial Intelligence systems is only suitable for simple diagnostic questions. | -3 | -1 | -2 | -1 | -1 |
| **41** | More effort should be put into the use of home measurements by patients. | 3 | 1 | -1 | 1 | 1 |
| **42** | High workload limits the implementation of innovation of diagnostics in primary care. | -2 | 0 | -2 | 0 | 3 |
| **43** | Digitization makes primary care more accessible and increases patient self-direction. | 1 | 1 | 1 | -2 | 1 |
| **44** | GPs need to collaborate more with data scientists and engineers to integrate Artificial Intelligence tools into primary care. | -1 | -1 | 2 | 1 | 0 |
| **45*** | Developers should collaborate more with healthcare providers and patients in the development of diagnostic innovations. | 1 | 2 | 0 | 2 | 1 |
| **46** | A different division of labor and way of working together in primary care is important for future-proof diagnostics in primary care. | -1 | 1 | -2 | 2 | 4 |
| **47** | Diagnostic innovation should ensure that patients can remain in primary care as much as possible. | 3 | -2 | 1 | -3 | -1 |
| **48** | Innovation should focus primarily on efficiency of the diagnostic process. | 0 | 0 | 3 | -2 | -1 |
| **49** | Artifical Intelligence should be used primarily to increase the efficiency of the diagnostic process. | -1 | 0 | 4 | -2 | 1 |
| **50** | There should be more focus on diagnostic innovation in general practitioner training. | 0 | 1 | 2 | 0 | 1 |
| **51** | An important factor in the success of a new diagnostic test is whether the health insurance company covers the cost of the test. | 0 | 2 | 3 | 1 | 0 |
| **52** | Diagnostic innovation should focus on cost savings. | -1 | -2 | 0 | -1 | -3 |
| **53** | The current funding system hinders innovation in the diagnostic process. | 1 | -1 | 0 | 3 | 0 |
| **54** | Digital remote diagnostics makes healthcare more accessible to patients. | 4 | 1 | -2 | -3 | -1 |
| **55** | Remote digital diagnostics causes inequities in access to care. | -4 | -2 | -3 | 2 | 0 |
| **56** | The interests of the patient must be at the center of the development and implementation of diagnostic innovations. | 0 | 4 | 2 | 1 | -1 |
| **57** | Technological developments such as Artificial Intelligence limit a physician's professional autonomy in making a diagnosis. | -3 | -3 | -4 | -4 | -3 |

**Consensus statements = statements that all perspectives agree or disagree with. These statements do not contribute to the distinction between the perspectives.*
