## Supplementary box 3 for "Perspectives on innovation of the diagnostic process in general practice: Q-methodological study"

**Box S3** Search strings for media

#### Nexis Uni

Search terms: Innovatie AND huisartsenpraktijk OR eerste lijn
Filters: last 5 years, content type: news. Group duplicates on.

#### Huisarts en Wetenschap

Search terms: innovatie diagnostiek
Filters: years 2019 – 2023

#### Google news

Search terms: innovatie AND huisarts
