## Supplementary box 2 for "Perspectives on innovation of the diagnostic process in general practice: Q-methodological study"

**Box S2** Search strings for specific journals

The specific journals were: The New England Journal of Medicine (NEJM) Catalyst, Healthcare: The Journal of Delivery Science and Innovation, and Journal of Medical Internet Research (JMIR). The first journal was searched manually, as it is not indexed in PubMed or Embase. Although the other two journals are included in the databases, our search was broad in scope. Therefore, we conducted an additional search within these journals to ensure relevant articles were not missed. These journals were searched to add other viewpoints to the search, including organizational, social, ethical, financial, and political viewpoints.

#### New England Journal of Medicine Catalyst Innovations in Care Delivery

title:'primary AND title:care' OR title:'general AND title:practice'

No filters applied, 35 results.

#### Healthcare: The Journal of Delivery Science and Innovation

Title: 'primary care' OR 'general practice' OR 'diagnostics' OR 'diagnose'

Results filtered for years 2018-2023, 36 results.

#### Journal of Medical Internet Research

Primary care [title/abstract/keyword] AND innovation [title/abstract/keyword] AND ('perspectives' OR 'qualitative' OR 'interview' OR 'perceptions' [title/abstract/keyword]) AND ('diagnosing' OR 'diagnostics' OR 'diagnose' OR 'diagnosis' [title/abstract/keyword])
