## Supplementary box 1 for "Perspectives on innovation of the diagnostic process in general practice: Q-methodological study"

**Box S1** Search strings for Pubmed and Embase

#### Pubmed

(((((((((((((((perspective[Title/Abstract]) OR (perspectives[Title/Abstract])) OR (opinion[Title/Abstract])) OR (opinions[Title/Abstract])) OR (attitudes[Title/Abstract])) OR (attitude[Title/Abstract])) OR (viewpoints[Title/Abstract])) OR (viewpoint[Title/Abstract])) OR (point of view[Title/Abstract])) OR (perception[Title/Abstract])) OR (belief[Title/Abstract])) OR (approach[Title/Abstract])) AND ((((((((((((((((((((doctor[Title/Abstract]) OR (doctors[Title/Abstract])) OR (physician[Title/Abstract])) OR (physicians[Title/Abstract])) OR (general practitioner[Title/Abstract])) OR (general practitioners[Title/Abstract])) OR (family doctor[Title/Abstract])) OR (gp[Title/Abstract])) OR (g.p.[Title/Abstract])) OR (family practitioner[Title/Abstract])) OR (family physician[Title/Abstract])) OR (nurse[Title/Abstract])) OR (family nurse[Title/Abstract])) OR (nurse practitioners[Title/Abstract])) OR (radiologist[Title/Abstract])) OR (radiologists[Title/Abstract])) OR (clinical chemist[Title/Abstract])) OR (clinical chemists[Title/Abstract])) OR (patient[Title/Abstract])) OR (patients[Title/Abstract]))) AND (((((((((((((diagnosis[Title/Abstract]) OR (diagnostics[Title/Abstract])) OR (examination[Title/Abstract])) OR (diagnosing[Title/Abstract])) OR (diagnostic imaging[Title/Abstract])) OR (point of care[Title/Abstract])) OR (point-of-care[Title/Abstract])) OR (point-of-care-testing[Title/Abstract])) OR (diagnostic test[Title/Abstract])) OR (diagnostic tests[Title/Abstract])) OR (diagnostic process[Title/Abstract])) OR (diagnostic tool[Title/Abstract])) OR (diagnostic tools[Title/Abstract]))) AND ((((((((((((((general practice[Title/Abstract]) OR (primary care[Title/Abstract])) OR (healthcare[Title/Abstract])) OR (primary healthcare[Title/Abstract])) OR (primary health care[Title/Abstract])) OR (outpatient care[Title/Abstract])) OR (family care[Title/Abstract])) OR (family health care[Title/Abstract])) OR (first-line care[Title/Abstract])) OR (first line care[Title/Abstract])) OR (first-line health care[Title/Abstract])) OR (first-line healthcare[Title/Abstract])) OR (first line health care[Title/Abstract])) OR (first line healthcare[Title/Abstract]))) AND (((qualitative[Title/Abstract]) OR (narrative[Title/Abstract])) OR (descriptive[Title/Abstract]))

Filters applied: full text, 2019-2023, humans, Language: Dutch/English.

975 results

#### Embase

('perspectives':ti,ab,kw OR 'perspective':ti,ab,kw OR 'opinion':ti,ab,kw OR 'opinions':ti,ab,kw OR 'attitude':ti,ab,kw OR 'attitudes':ti,ab,kw OR 'viewpoints':ti,ab,kw OR 'viewpoint':ti,ab,kw OR 'views':ti,ab,kw OR 'view':ti,ab,kw OR 'point of view':ti,ab,kw OR 'perception':ti,ab,kw OR 'belief':ti,ab,kw OR 'approach':ti,ab,kw)

AND

('doctor':ti,ab,kw OR 'doctors':ti,ab,kw OR 'physician':ti,ab,kw OR 'physicians':ti,ab,kw OR 'general practitioner':ti,ab,kw OR 'general practitioners':ti,ab,kw OR 'family doctor':ti,ab,kw OR 'gp':ti,ab,kw OR 'g.p.':ti,ab,kw OR 'family practitioner':ti,ab,kw OR 'family physician':ti,ab,kw OR 'nurse':ti,ab,kw OR 'family nurse':ti,ab,kw OR 'nurse practitioner':ti,ab,kw OR 'radiologist':ti,ab,kw OR 'clinical chemist':ti,ab,kw OR 'clinical chemists':ti,ab,kw OR 'patient':ti,ab,kw OR 'patients':ti,ab,kw)

AND

('diagnosis':ti,ab,kw OR 'diagnostics':ti,ab,kw OR 'examination':ti,ab,kw OR 'diagnosing':ti,ab,kw OR 'diagnostic process':ti,ab,kw OR 'diagnostic imaging':ti,ab,kw OR 'point of care':ti,ab,kw OR 'point-of-care':ti,ab,kw OR 'point-of-care testing':ti,ab,kw OR 'diagnostic test':ti,ab,kw OR 'diagnostic tests':ti,ab,kw OR 'diagnostic tool':ti,ab,kw OR 'diagnostic tools':ti,ab,kw)

AND

('innovation':ti,ab,kw OR 'innovative':ti,ab,kw OR 'new':ti,ab,kw OR 'improving':ti,ab,kw OR 'improvement':ti,ab,kw OR 'improved':ti,ab,kw OR 'improvements':ti,ab,kw OR 'invention':ti,ab,kw OR 'inventions':ti,ab,kw OR 'inventive':ti,ab,kw OR 'innovational':ti,ab,kw)

AND

('general practice':ti,ab,kw OR 'primary care':ti,ab,kw OR 'primary healthcare':ti,ab,kw OR 'primary health care':ti,ab,kw OR 'outpatient care':ti,ab,kw OR 'family care':ti,ab,kw OR 'family health care':ti,ab,kw OR 'first-line care':ti,ab,kw OR 'first line care':ti,ab,kw OR 'first-line health care':ti,ab,kw OR 'first line health care':ti,ab,kw OR 'first line healthcare':ti,ab,kw)

AND

('qualitative':ti,ab,kw OR 'narrative':ti,ab,kw OR 'descriptive':ti,ab,kw)

AND ([dutch]/lim OR [english]/lim) AND [humans]/lim AND [2013-2023]/py

Results: 576
