## Supplementary table 1 for "Perspectives on innovation of the diagnostic process in general practice: Q-methodological study"

| **Table S1** Search terminology |  |
| --- | --- |
| **Search criteria** | **Terminology** |
| Perspectives | ‘perspective’, ‘perspectives’, ‘opinion’, ‘opinions’, ‘attitudes’, ‘attitude’, ‘viewpoint’, ‘viewpoints’, ‘views’, ‘view’, ‘point of view’, ‘perception’, ‘belief’, ‘approach’ |
| Multidisciplinary stakeholders | ‘doctor’, ‘doctors’, ‘physician’, ‘physicians’, ‘general practitioner’, ‘general practitioners’, ‘family doctor’, ‘GP’, ‘G.P.’, ‘family practitioner’, ‘family physician’, ‘nurse’, ‘family nurse’, ‘nurse practitioners’, ‘radiologist’, ‘radiologists’, ‘clinical chemist’, ‘clinical chemists’, ‘patient’, ‘patients’ |
| Diagnostics | ‘diagnosis’, ‘diagnostics’, ‘examination’, ‘diagnosing’, ‘diagnostic imaging’, ‘point of care’, ‘point-of-care’, ‘point of care testing’, ‘diagnostic test’, ‘diagnostic tests’, ‘diagnostic process’, ‘diagnostic tool’, ‘diagnostic tools’ |
| Innovation | ‘innovation, ‘innovative’, ‘new’, ‘improving’, ‘improved’, ‘improvements, ‘improvement’, ‘invention’, ‘inventions’, ‘inventive’, ‘innovational’ |
| General practice domain | ‘General practice’, ‘primary care’, ‘primary healthcare’, ‘primary health care’, ‘outpatient care’, ‘family care’, ‘family health care’, ‘first-line care’, ‘first line care’, ‘first-line health care’, ‘first-line healthcare’, ‘first line health care’, ‘first line healthcare’ |
| Qualitative | ‘qualtitative’, ‘narrative’, ‘descriptive’ |
